# Deep Learning Frame Prediction for Abbreviated Low-Dose Dynamic PET Protocols on the PennPET Explorer

**DOI:** 10.64898/2026.08.25.26361357

**Authors:** Joran Courtens, Florence M. Muller, Elizabeth J. Li, Christian Vanhove, Stefaan Vandenberghe, Austin R. Pantel, Joel S. Karp, Margaret E. Daube-Witherspoon

**Affiliations:** Faculty of Engineering and Architecture, Ghent University, Ghent, Belgium; Department of Radiology, University of Pennsylvania, Philadelphia PA, United States

**Author notes:** Corresponding author: Florence M Muller. Authors JC and FMM contributed equally to this work.

**Keywords:** Abbreviated Dynamic Protocols, Deep Learning, Kinetic Modelling, Long Axial Field-of-View PET

## Abstract

Dynamic positron emission tomography (PET) with long axial field-of-view (LAFOV) scanners enables multi-organ imaging and kinetic quantification beyond static (late-phase) imaging; however, the long times typically required for dynamic acquisitions remain clinically impractical. This study evaluates a deep learning (DL) framework to enable abbreviated dynamic PET acquisitions, comparing single-time-window (STW, early dynamic data only) and dual-time-window (DTW, early dynamic data plus a late 5-min static frame) protocols with early dynamic scan durations of 5-30 min and dose levels ranging from 360 MBq to 18 MBq. Seventeen 60-min dynamic [^18^F]FDG datasets were first motion-corrected using a staggered FALCON pipeline and then used to train and test a spatiotemporal DL model for autoregressive frame prediction. Performance was assessed across the full quantitative workflow, from DL-predicted frames and time-activity curves to organ-based kinetic modeling and voxel-wise parametric imaging in multiple tissues and two patient cohorts. DTW protocols consistently outperformed STW, better preserving late-phase kinetics. For a 15-min early dynamic scan, adding a late 5-min scan reduced mean absolute K_i_ difference from 23% (STW) to 17% (DTW) in the liver and from 26% to 15% in the thalamus. DTW + DL further reduced errors to ≤10% in the liver, thalamus, and breast lesion, and 16% in muscle. Our recommended protocol, 15-min early dynamic scan plus a 5-min late scan with DL, remained robust to up to a 5-fold dose reduction (∼74 MBq). Overall, these findings support DL-enabled abbreviated, low-dose dynamic LAFOV PET as a clinically feasible approach for accurate kinetic quantification.

## I. INTRODUCTION

Dynamic positron emission tomography (PET) provides temporal information on tracer uptake, enabling estimation of kinetic parameters that offer insights into physiological processes beyond conventional static imaging [1, 2]. Clinical PET primarily relies on the standardized uptake value (SUV), a semi-quantitative metric measured from a static image acquired at a fixed time after tracer injection (e.g., 60 min for [^18^F]FDG) [3, 4]. Although widely used, SUV only captures the tracer uptake at a single time point, which may be changing at the time of imaging, depending on the tracer kinetics [5]. Dynamic PET models tracer delivery, transport, and retention using compartmental modelling [1] or graphical approaches (e.g., Patlak analysis [6]), enabling the separation of specific from non-specific uptake and the estimation of multiple kinetic parameters (e.g., blood flow, tracer delay, metabolic flux) that improve disease characterization and therapy response assessment [1, 7-10].

On standard axial field-of-view (SAFOV) PET scanners, dynamic imaging is generally limited to a single bed position, restricting anatomical coverage and preventing image-derived input function (IDIF) extraction when major blood pools lie outside the AFOV. Although continuous bed motion protocols partially address these limitations [11], they remain constrained by the sparse temporal sampling and lower sensitivity of SAFOV systems. In contrast, long AFOV (LAFOV) scanners (AFOV > 60-70 cm) enable simultaneous imaging of most or all of the body with up to 3-fold higher sensitivity per organ [12], facilitating multi-organ interaction studies [13], reducing image noise, and improving kinetic parameter estimation [14]. This sensitivity boost also enables reliable reconstruction of very short dynamic frames (e.g. ≤5s) for IDIF extraction [1, 15]. Despite these benefits, routine dynamic LAFOV PET remains clinically impractical because imaging over the full uptake period is time-consuming (typically ≥60 min for [^18^F]FDG) and disrupts clinical workflow. Moreover, prolonged acquisitions increase susceptibility to patient motion, which can impact downstream kinetic analysis [16].

To facilitate clinical translation, abbreviated dynamic scan protocols have been proposed for LAFOV PET. For example, dynamic [^18^F]FDG scans have been shortened from 60-75 to 30-45 min while retaining acceptable parameter estimates for K_i_ and K_1_ in tumors and normal organs [17, 18]. Accurate kinetic quantification of lung tumors has also been achieved using a 26-min dynamic [^18^F]FAPI PET acquisition instead of the conventional 60-min protocol [19]. Dual-time-window protocols, which combine an early dynamic scan with a late static scan (∼60 min post-injection (p.i.)) to preserve SUV reporting, have also been studied. Prior work [20-22] has demonstrated accurate estimation of the FDG influx rate (K_i_) using two short dynamic scans, or a dynamic plus a static scan. For example, Viswanath et al. [21] found K_i_ bias within ±10% for breast tumors using an abbreviated protocol combining a dynamic scan (0-15 min) plus a static scan (60-65 min). However, these protocols require two separate imaging sessions, each with a CT for attenuation correction, as well as inter-scan image registration for subsequent kinetic analysis. Alternatively, population-based input function methods replace the early scan by estimating the blood input curve from a reference population and scaling it using late image-derived measurements. These methods have enabled accurate Patlak and compartmental modelling from abbreviated late scans (e.g., 30-60 or 40-60 min p.i. for [^18^F]FDG on LAFOV PET) [23, 24], but remain susceptible to inter-subject variability.

Recent advances in deep learning (DL) have created new opportunities to shorten dynamic PET scans. Existing DL approaches have focused on predicting missing dynamic frames [25-28], estimating arterial input functions [29], or directly generating kinetic parameters and parametric images [30-32]. Several methods further incorporate kinetic models as physiological constraints [26, 30, 32]. For LAFOV PET, Yang et al. [25] introduced a bidirectional CycleGAN to reconstruct 1-h dynamic sequences from early (0-10 min) and late (50-60 min) acquisitions for [^68^Ga]PSMA and [^68^Ga]FAPI PET. Gao et al. [32] extended this GAN-based framework by integrating a parametric fitting module. Wen et al. [31] developed a diffusion model that directly generates Patlak K_i_ images from abbreviated [^18^F]FDG acquisitions, reporting that late (50-60 min) single-time-window protocols generally outperformed early (10-20 min) acquisitions, except for brain regions where uptake is higher due to less dephosphorylation relative to the rest of the body. Most closely related to this work is the study by Ding et al. [28] comparing multiple abbreviated protocols with early dynamic durations of 10, 15, 20, and 25 mins, optionally supplemented with a 5-min late static scan. Using a bidirectional generative network (similar to [25]) with convolutional recurrent neural network and long short-term memory (LSTM) cells, they showed accurate [^18^F]FDG kinetic quantification from a 10-min early scan with a 5-min late scan for patients with breast cancer or pulmonary nodules.

While these studies demonstrate the potential of DL to enable abbreviated dynamic LAFOV PET imaging, only Ding et al. evaluated their DL frame prediction model across the complete quantitative pipeline, from frame image quality to organ- and voxel-based kinetic quantification. In parallel, growing interest in same-session multi-tracer PET has increased the need for dose reduction to limit radiation exposure. Dynamic dual-tracer PET enables multiple physiological pathways to be assessed in a single imaging session using low-/high-dose injection paradigms for the two tracers [33-35]. However, lower dose scans have reduced count statistics, increasing noise in dynamic PET frames and time-activity curves (TACs), which degrades the accuracy and precision of kinetic parameter estimation and propagates noise into parametric images [36]. These challenges are further exacerbated when combined with abbreviated scan protocols.

The work presented here extends previous developments and analyses of DL methods for abbreviated dynamic LAFOV PET. In particular, a spatiotemporal neural network combining an encoder with ConvLSTM cells [37] and a CNN decoder was developed for autoregressive prediction of missing (unacquired) dynamic [^18^F]FDG PET frames. The DL networks either predicted subsequent frames following an early dynamic scan or frames within a temporal gap between two scans for single- and dual-time-window protocols, respectively, and was evaluated for early dynamic scan durations of 5-30 min and dose levels ranging from ∼ 360 MBq (full dose) down to ∼ 18 MBq (5% dose). An important aspect to the quantitative evaluation is that motion correction preprocessing is implemented to ensure alignment of paired abbreviated and full-sequence dynamic PET data prior to supervised training. Performance was assessed through the complete quantitative pipeline, from reconstructed frames and TACs to VOI-based kinetic modeling and parametric imaging, in multiple tissues and two independent patient cohorts. By jointly investigating scan duration and dose, this study aims to identify the shortest, lowest-dose dynamic LAFOV PET scan protocol that preserves quantitative accuracy.

## II. Materials and Methods

### A. Data Acquisitions and Image Reconstruction

All dynamic data were acquired on the PennPET Explorer [38], a 142-cm LAFOV PET system. Seventeen 60-min dynamic [^18^F]FDG PET scans (359 ± 41 MBq; BMI: 29.0 ± 7.7 [range: 20.7-43.1] kg/m^2^) were obtained from two research protocols. (**1**) Thirteen datasets were from a alcohol use disorder (AUD) study investigating the pharmacokinetic effects of nutritional ketone ester (KE) on brain glucose and ketone metabolism [39]. (**2**) Four additional FDG datasets were of patients with breast cancer (two estrogen receptor-positive, ER+, and two with triple-negative breast cancer, TNBC) [35]. Each 60-min list-mode data was parsed into 39 frames: 12x 5s, 6x 10s, 3x 20s, 2x 30s, 6x 1 min, and 10x 5 min. Frames were reconstructed in 4×4×4 mm^3^ voxels using a time-of-flight (TOF) ordered-subsets expectation-maximization [40, 41] algorithm with default reconstruction parameters and the following corrections: CT-based attenuation, normalization, delay-based randoms, and TOF-SSS (single scatter simulation) scatter estimation [42].

Additionally, to emulate low-dose dynamic [^18^F]FDG PET studies, each frame was subsampled to 1/2, 1/5, 1/10, and 1/20 of the original counts with event shuffling and splitting. The scatter and randoms correction files were proportionally scaled from the full-dose data to each subsampled frame.

### B. Dynamic Scan Protocol Abbreviations

Two abbreviated protocols were studied by retrospectively processing portions of the acquired 60-min dynamic list-mode data: *single-time-window* (STW) and *dual-time-window* (DTW) protocols (**Fig. 1**). The STW protocol used early dynamic data only, starting at injection and continuing until an early cut-off time t_early-dyn_. Six abbreviated scan times were considered, with t_early-dyn_ of 5, 10, 15, 20, 25, and 30 mins. The DTW protocol used the same early dynamic data (= 0-t_early-dyn_ mins) supplemented with a late 5-min static frame (55-60 min p.i.), enabling SUV measurement for clinical interpretation. Early scan durations of t_early-dyn_ < 5 mins were considered insufficient for reliable whole-body FDG influx (K_i_) estimation.

**Fig. 1:**
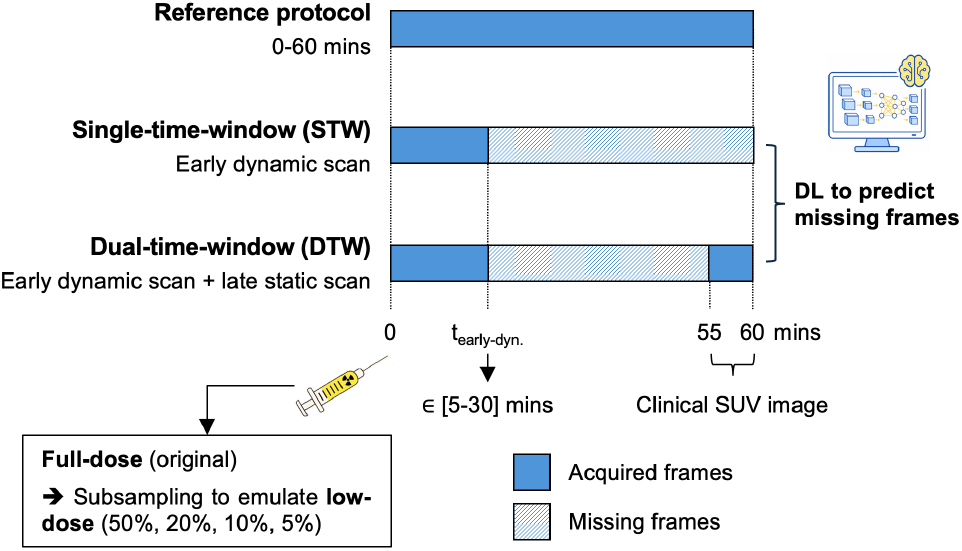
Overview of the abbreviated dynamic scan protocols. The single-time-window (STW) protocol uses only early dynamic data (0-t_early-dyn_). The dual-time-window (DTW) protocol combines the early dynamic data with a 5-min late scan (55-60 mins p.i.). A DL model aims to predict the missing dynamic frames between t_early-dyn_ - 60 mins for STW, or between t_early-dyn_ - 55 mins for DTW.

### C. Preprocessing: Motion Correction

Motion correction (MoCorr) was performed as a preprocessing step to co-register all dynamic frames of each study, ensuring voxel-level spatial alignment between the abbreviated sequence (i.e., network input: early dynamic frames with an optional late static frame) and the 60-min dynamic sequence (i.e., network target containing the missing frames). This allowed studies affected by patient motion to be reliably included in supervised DL model training. MoCorr was applied to all 17 full-dose 60-min dynamic [^18^F]FDG PET studies using the open-source FALCON V2 tools, an established registration framework for dynamic PET [16]. The resulting deformation fields were then applied to the corresponding subsampled datasets.

Two FALCON-based MoCorr pipelines were implemented: the default FALCON (**Fig. 2a**) and a subject-specific staggered FALCON (**Fig. 2b**). Additional comparisons are provided in the **Supplemental Material** (**Fig. S1-S4**). All results in this manuscript are based on datasets corrected with the staggered FALCON pipeline, which was implemented to address two limitations of the default pipeline [16]: (1) residual respiratory motion in the early dynamic frames (0-90s p.i.) and (2) registration inaccuracies caused by bladder-filling differences between early and late frames. Staggered FALCON groups dynamic frames into sequential batches of comparable tracer distributions. Starting from the last frame (55-60 min p.i.) as a fixed reference, registration is performed within each batch, with each newly motion-corrected frame serving as reference for the preceding frame in the next batch.

**Fig. 2:**
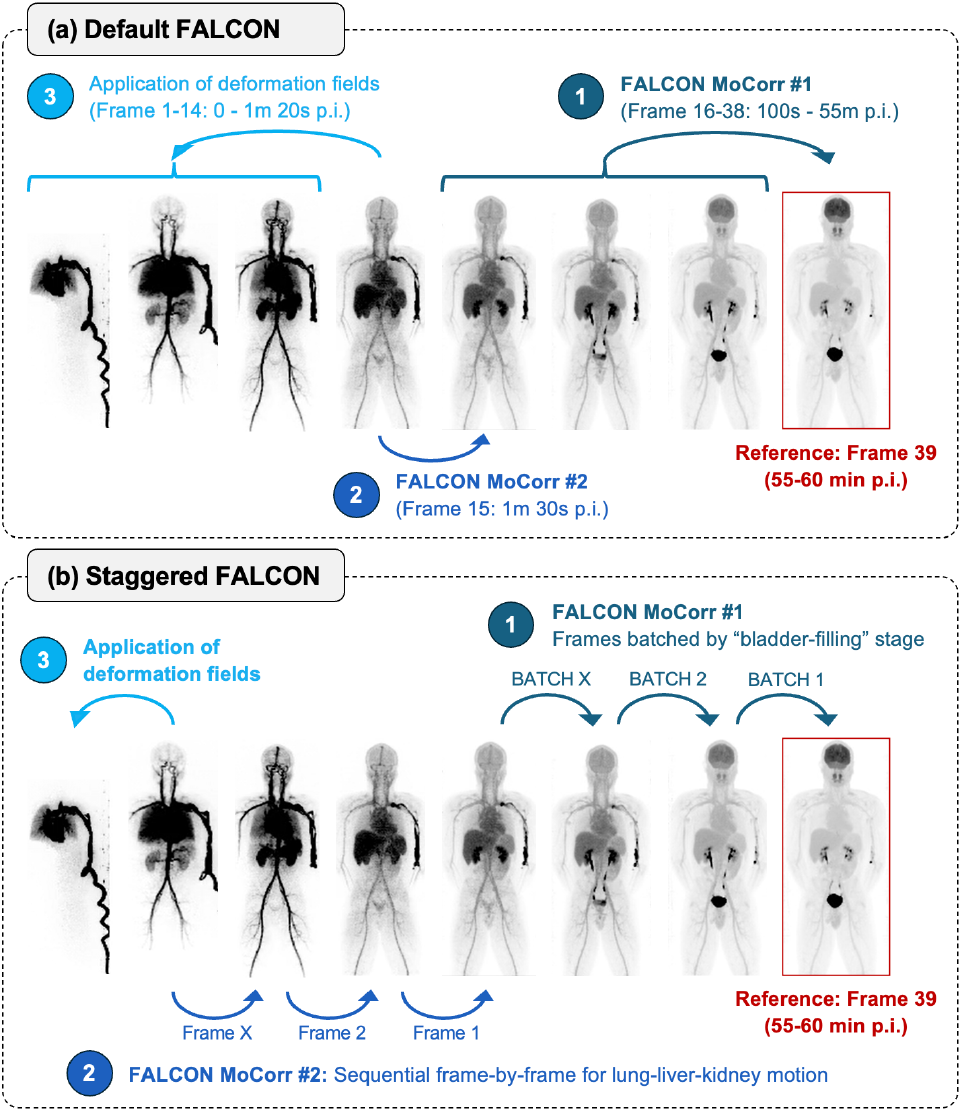
Two FALCON-based motion correction (MoCorr) pipelines. **(a) Default FALCON**: MoCorr Step #1 performs registration of frames 16-38 (1.5-55 min p.i.) to the reference frame (55-60 min p.i.). MoCorr Step #2 then registers frame 15 (80-90s p.i.) to the motion-corrected frame 16, with resulting deformation fields subsequently applied to align all earlier frames 1-14 (0-80s p.i.). **(b) Staggered FALCON**: Dynamic frames are grouped into batches with comparable tracer distribution. Registration is performed within each batch and repeated across batches, with each newly motion-corrected frame serving as the reference for the preceding batch. For early dynamic frames (50-90s p.i.), each frame is treated as individual batch, resulting in sequential frame-by-frame registration.

### D. Deep Learning Network Architecture

We propose and test a spatiotemporal neural network that combines a ConvLSTM-based encoder with a CNN decoder in a U-Net architecture for autoregressive prediction of missing dynamic PET frames (**Fig. 3**). Given an abbreviated sequence of early frames (and an additional 5-min late frame for DTW), the neural network recursively predicts each subsequent missing frame, aiming to complete the 60-min sequence.

**Fig. 3:**
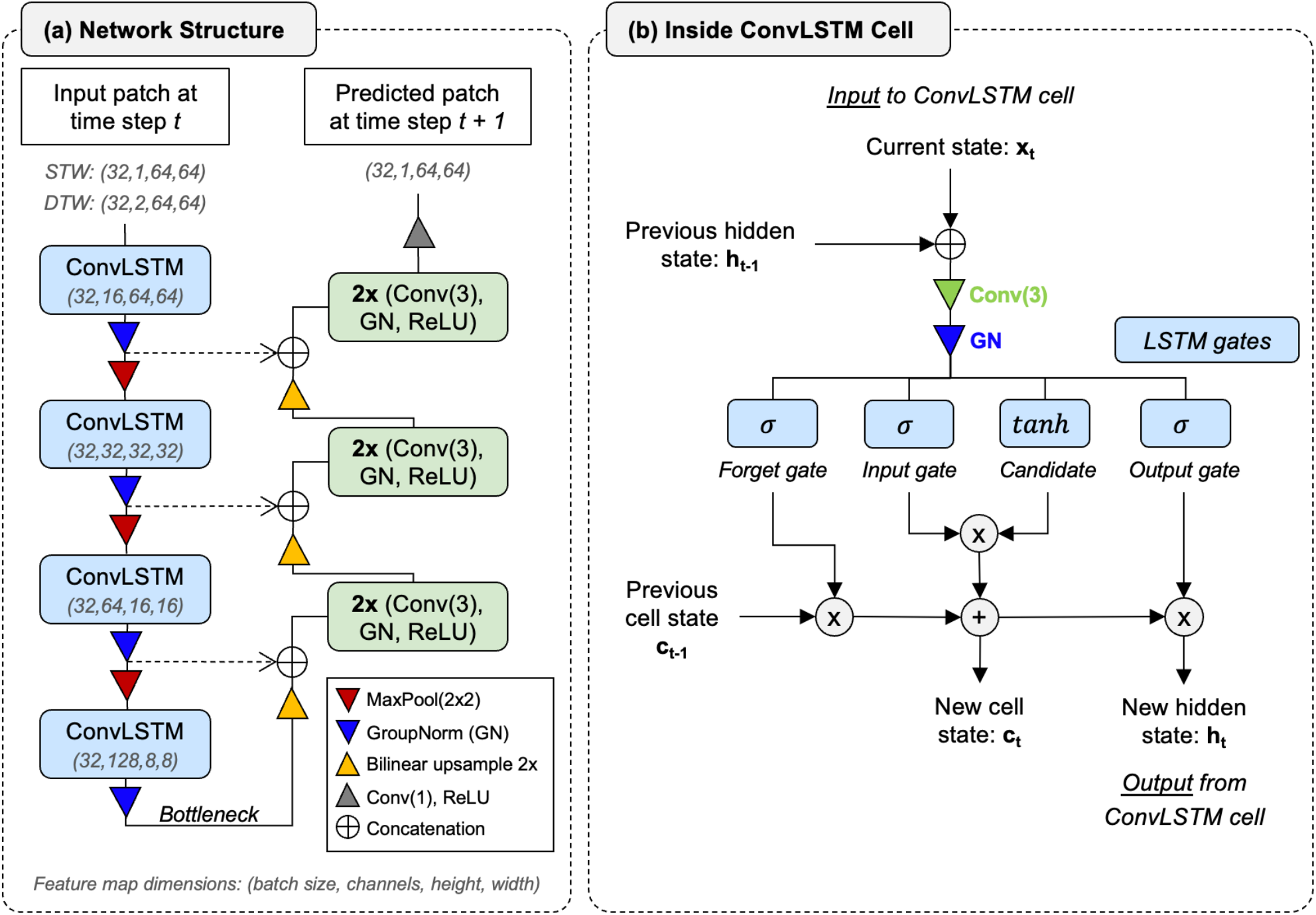
Proposed ConvLSTM-CNN architecture for autoregressive dynamic frame prediction. **(a)** 2D U-Net network with a ConvLSTM encoder and CNN decoder. Predicted frame at time step t+1 is fed back into the network to predict subsequent frames. Gray numbers (in parentheses) denote the feature map dimensions in the format (batch size, channels, height, width) with STW and DTW indicating the corresponding input patch dimensions. **(b)** Internal structure of a ConvLSTM cell, showing the convolution, group normalization (GN), and LSTM gating mechanism used to update the cell and hidden states.

The encoder processes the input sequence one frame at a time using a hierarchy of ConvLSTM cells [37] operating with 16, 32, 64, and 128 feature channels. At each level, the ConvLSTM cell receives the current feature map (x_t_) and previous hidden state (h_t-1_) as input. A 3×3 convolution followed by group normalization (GN) is applied before updating the cell state (c_t_) and hidden state (h_t_) through the standard LSTM gating mechanism, consisting of an input gate, forget gate, output gate (all using sigmoid activations), and a candidate memory state (using a hyperbolic tangent activation). The updated hidden state (h_t_) is passed through a GN, retained as a skip connection, and down-sampled by 2×2 max-pooling before being passed to the next encoder level. At the bottleneck, the encoder operates on 8×8 feature maps with 128 channels.

From the bottleneck, the decoder generates the predicted frame for time step t+1 through three up-sampling stages. Each stage consists of bilinear interpolation, concatenation with the corresponding encoder skip connection, and a double 3×3 convolution block with GN and ReLU activation. A final 1×1 convolution and ReLU activation convert the 16-channel feature map to a single-channel output corresponding to the predicted (patch) frame for the next time step (= x_t+1_). The network contains around 1.4 million trainable parameters.

The predicted frame (x_t+1_) is fed back into the network as input for the next autoregressive step, enabling recursive prediction of subsequent frames. For the DTW protocol, the 5-min late frame is concatenated with the current input frame along the channel dimension at every autoregressive time step.

### E. Training Procedure and Inference Network Architecture

The 17 studies were split into training (9), validation (2), and test (6) cohorts. Training and validation data were drawn from the AUD/KE study, while the test cohort included two subjects with AUD (baseline and post-KE intervention) and all four breast cancer subjects. Mixed-dose abbreviated data from five dose levels (full, 1/2, 1/5, 1/10, 1/20) were used as network input and mapped to the corresponding 0-60 min full-dose reference. For subsampling, list-mode data were shuffled and split to generate statistically independent low-count replicates, yielding two, five, ten, and ten images for the 1/2-, 1/5-, 1/10- and 1/20-count data, respectively, resulting in 28 input datasets per study. Networks were trained using 64×64 voxel patches extracted from 2D coronal and sagittal slices. Slices were only retained if they contained at least one valid patch location, resulting in 49,883 paired input-target slice entries across all count-levels and slices. Candidate patch centers were defined by voxel values exceeding 15% of the mean slice intensity. Valid patch coordinates were precomputed, and each slice contributed one randomly selected patch per training epoch.

A total of 12 models were trained, corresponding to each t_early-dyn_ configuration for both the STW and DTW protocols. Networks were optimized using Adam (initial learning rate 1.0E-4, with exponential decay of 0.95 per epoch, and batch size 32) with a voxel-wise L1 loss. For DTW, an additional weighted penalty was applied to the predicted final frame, giving *L* = *L*_*Missing*_ + 0.05 · *L*_*Last*_, where *L*_*Missing*_ and *L*_*Last*_ denote the L1 losses for the missing frames and final frame (frame 39 at 55-60 min p.i.), respectively. Scheduled teacher-forcing [43] was used to mitigate error accumulation during autoregressive prediction, with the probability of substituting target patches for predictions decreasing from 50% at epoch 0 to 0% by epoch 50. Training was stopped early when the validation loss did not improve for five consecutive epochs.

Inference used overlapping 64×64 patches extracted from coronal slices using a sliding-window strategy with a stride of 32 (50% overlap between adjacent patches). A 2D Hanning window was applied before patch aggregation to suppress boundary artifacts. Predicted coronal slices were stacked into 3D dynamic frames and combined with the acquired early frames and, for DTW, with the late reference frame.

### F. Performance Evaluation

The performance of the DL-based frame prediction network was first evaluated using full-dose data by comparing STW and DTW protocols. The best-performing abbreviated scan protocol was then evaluated at 50%, 20%, 10%, and 5% count levels to assess robustness under low-dose conditions. Volumes-of-interest (VOIs) were delineated on the full-dose motion-corrected data using PMOD V4.5 (Bruker Switzerland AG, Zurich, Switzerland), and then applied to the abbreviated and count-subsampled datasets. TACs were extracted for the liver, psoas muscle, thalamus, and breast lesions, and fitted using an irreversible two-tissue-compartment model (2-TCM, k_4_=0). The liver was modelled using a dual-blood input function (IF) to account for both the hepatic arterial and portal venous blood supply [44]. An IDIF was obtained from the descending aorta (DA) for the other organs, with blood delay and blood volume also fitted. The kinetic parameter of interest for this work was the net influx rate K_i_ [45], determined by a 2-TCM and by voxel-wise Patlak K_i_ parametric images [6].

## III. Results

The MoCorr results are presented in the **Supplemental Material**, showing that the staggered FALCON approach improve image alignment and reduce registration artifacts compared with default FALCON (**Fig. S1-S2**), while both methods achieved comparable correction of gross body motion (**Fig. S3**). Frame-by-frame normalized correlation coefficient (NCC) (**Fig. S4**) was consistently higher after MoCorr, with staggered FALCON increasing NCC by 33% relative to no MoCorr, versus a 24% improvement for default FALCON.

### A. STW vs. DTW Protocols

Image quality improved consistently with longer t_early-dyn_, as shown by the difference images of the DL-predicted frames (50-55 min p.i., **Fig. 4**). Across all scan durations, DTW showed closer visual agreement with the reference than STW, with the greatest difference at 5-min: STW was blurred and failed to recover the bladder (**Fig. 4**, arrow). Quantitative metrics supported these observations: DTW + DL consistently achieved lower NRMSE and higher SSIM than STW + DL.

**Fig. 4:**
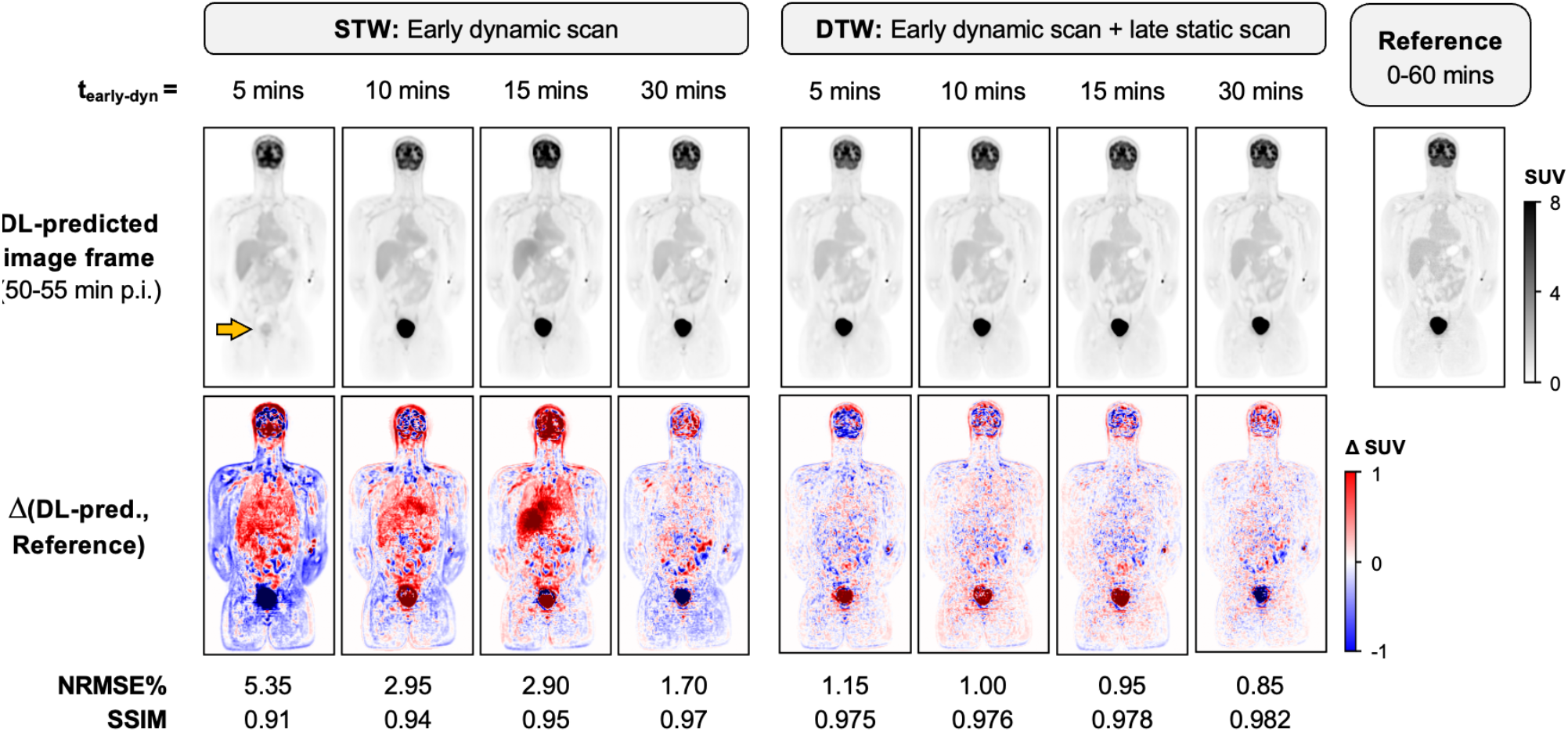
DL-predicted image frames at 50-55 min p.i. shown for a representative subject from the AUD/KE FDG study, with voxel-wise difference images (ΔSUV) relative to the corresponding measured full-dose reference frame (344 MBq [^18^F]FDG). Results are presented for STW and DTW protocols with different early scan durations (t_early-dyn_), together with the corresponding NRMSE% and SSIM values. The orange arrow highlights the absent bladder in the DL-predicted image from 5-min STW.

**Fig. 5** compares tissue TACs derived from the DL-predicted dynamic sequences with the 60-min reference for STW and DTW protocols with t_early-dyn_ of 5, 10, 15, and 30 mins. Lung TACs agreed well with the reference across all abbreviated protocols, likely because lung activity was already in a stable washout phase after the initial peak, facilitating network predictions from limited early data. Larger differences were observed in the liver, thalamus, muscle, and breast lesion, where DTW + DL consistently reproduced late-phase kinetics more accurately than STW + DL. This improvement was most evident for breast lesions: STW predicted a continuous decline in activity, whereas DTW better captured the increasing (TNBC) or stable (ER+) uptake seen in the reference.

**Fig. 5:**
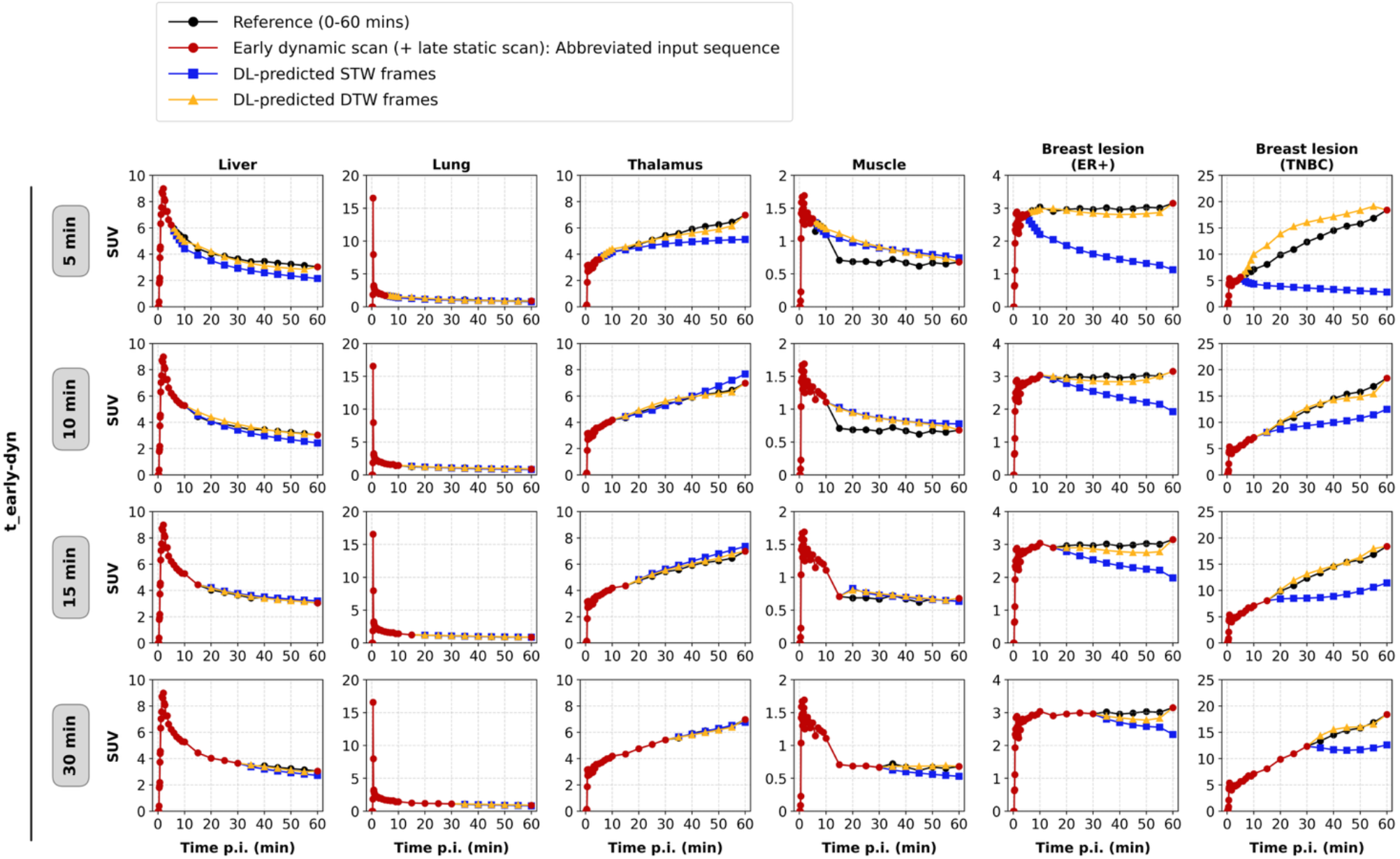
Representative TACs extracted from the liver, lung, thalamus, muscle, and a breast lesion in an ER+ breast cancer subject and from a TNBC subject. The reference TACs acquired over the 0-60 min dynamic scan are shown in black. The abbreviated early dynamic scan available as network input is shown in red, including the last frame (late static scan) for the DTW protocol. The DL-predicted image sequences for STW (blue) and DTW (orange) protocols are shown for early scan durations (t_early-dyn_) of 5, 10, 15, and 30 mins. Note, the y-axis scales differ across tissues, particularly for the two lesion types.

**Fig. 6** reports the mean absolute percent difference in FDG K_i_ from the 2-TCM fitting for STW and DTW with DL-based frame prediction, as well as without DL to illustrate the impact of DL on these protocols. Errors and inter-subject variability decreased with longer t_early-dyn_, with the largest differences for t_early-dyn_ below 10 min. Adding the late 5-min scan (DTW) improved accuracy even without DL; for example, for t_early-dyn_ = 15 min, the mean absolute percent K_i_ difference decreased from 23% (STW) to 17% (DTW) in the liver and from 26% to 15% in the thalamus. DTW + DL further reduced mean errors, reaching <10% in liver and <7% in thalamus. Muscle, a low-flux region (with reference K_i_: 0.17-0.50 ml.min^-1^.100cm^-3^), showed the largest errors across all protocols (16% at t_early-dyn_ = 15 min) and highest inter-subject variability among tissues. For the breast lesion, DTW + DL consistently outperformed STW + DL, achieving errors within 10% for t_early-dyn_ ≥ 15 mins. **Table I** summarizes the tissue-specific protocol recommendations for the shortest scan duration required to achieve an absolute FDG K_i_ difference of <10% relative to the 0-60 min reference.

**Fig. 6:**
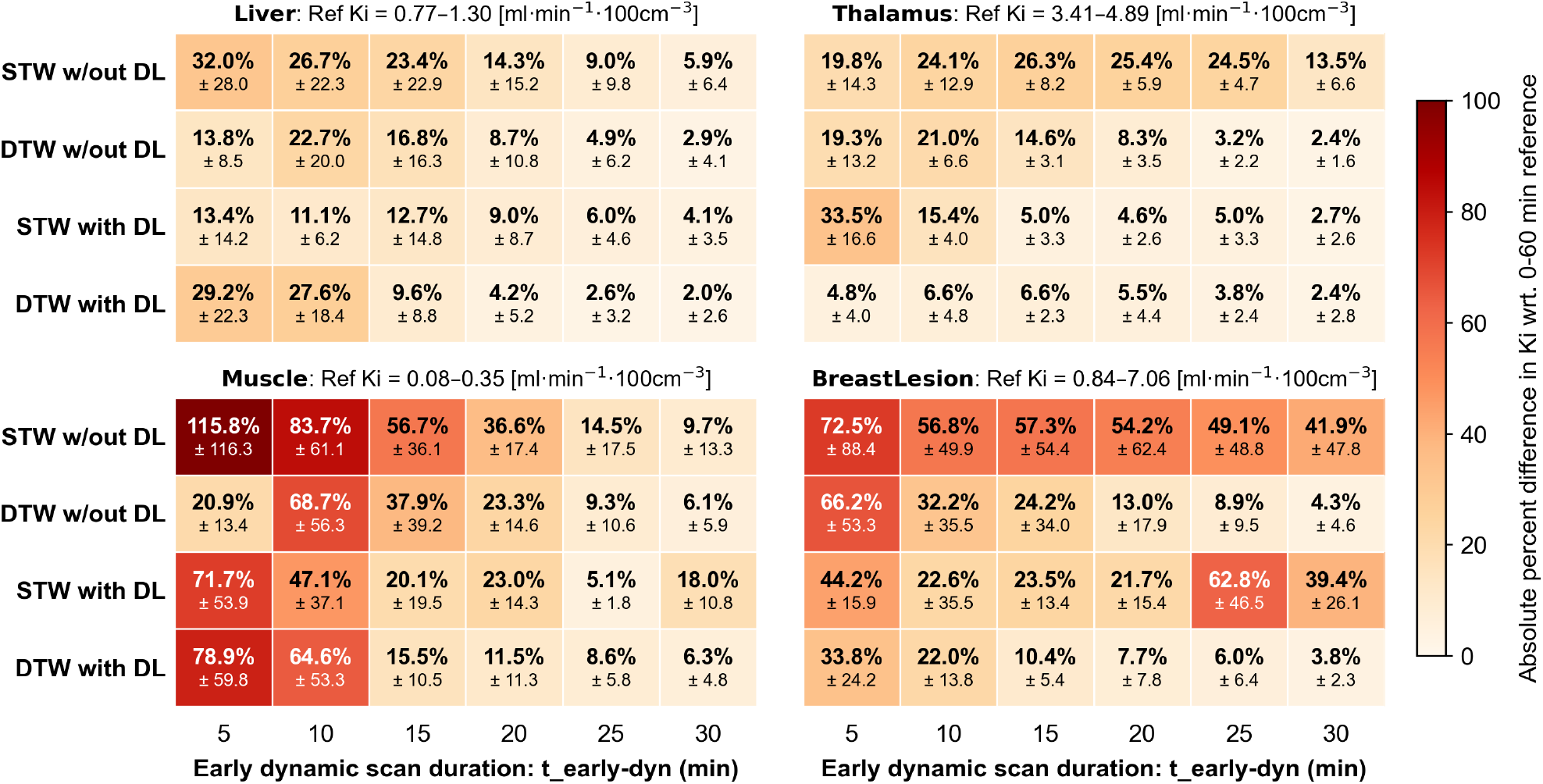
Mean absolute percent difference in FDG K_i_ (± standard deviation) relative to the 0-60 min dynamic reference scan for the liver, thalamus, muscle, and breast lesion. Results are averaged across six test subjects, except for the breast lesion VOI, which includes four test subjects. Each heatmap summarizes the performance of the four abbreviated protocols at different early dynamic scan durations: STW without DL, DTW without DL, STW with DL, and DTW with DL. The K_i_ values displayed in the subtitles indicate the range (min-max) of reference K_i_ values from the 60-min scan across testing subjects for a particular region of interest.

**TABLE I:** Shortest scan duration (in mins) to achieve an absolute percent difference in FDG K_i_<10% relative to 0-60 min reference K_i_. Values represent the average across test subjects of the earliest scan duration at which this bias remained below the 10% threshold for all subsequent longer scan durations.

|  | STW<br>w/out DL | DTW<br>w/out DL | STW<br>with DL | DTW<br>with DL |
| --- | --- | --- | --- | --- |
| **Liver** | 24.1 | 19.2 | 18.6 | 14.9 |
| **Thalamus** | > 30 | 18.6 | 12.6 | 8.6 |
| **Muscle** | 29.7 | 24.8 | > 30 | 22.6 |
| **Lesion** | > 30 | 23.7 | > 30 | 15.8 |

**Fig. 7** compares Patlak-derived K_i_ images of a test subject with high myocardial net influx. Non-DL STW produced severely degraded K_i_ maps at short scan durations, whereas non-DL DTW preserved the myocardial walls but remained noisy and overestimated influx values for t_early-dyn_ < 15 min. With DL, noise in the K_i_ maps was reduced for both protocols. However, for the STW + DL protocol, the myocardium was absent for t_early-dyn_ of 5 and 10 min, and showed an acceptable agreement with the reference (SSIM > 0.85, NRMSE% < 10%) only for t_early-dyn_ of 30 min. In contrast, DL-predicted DTW closely matched the reference across all t_early-dyn_, including 5-min. Similar trends were observed in brain and lesion K_i_ maps: DTW consistently preserved morphology and contrast, while STW frequently underestimated or failed to recover structures at shorter scan durations.

**Fig. 7:**
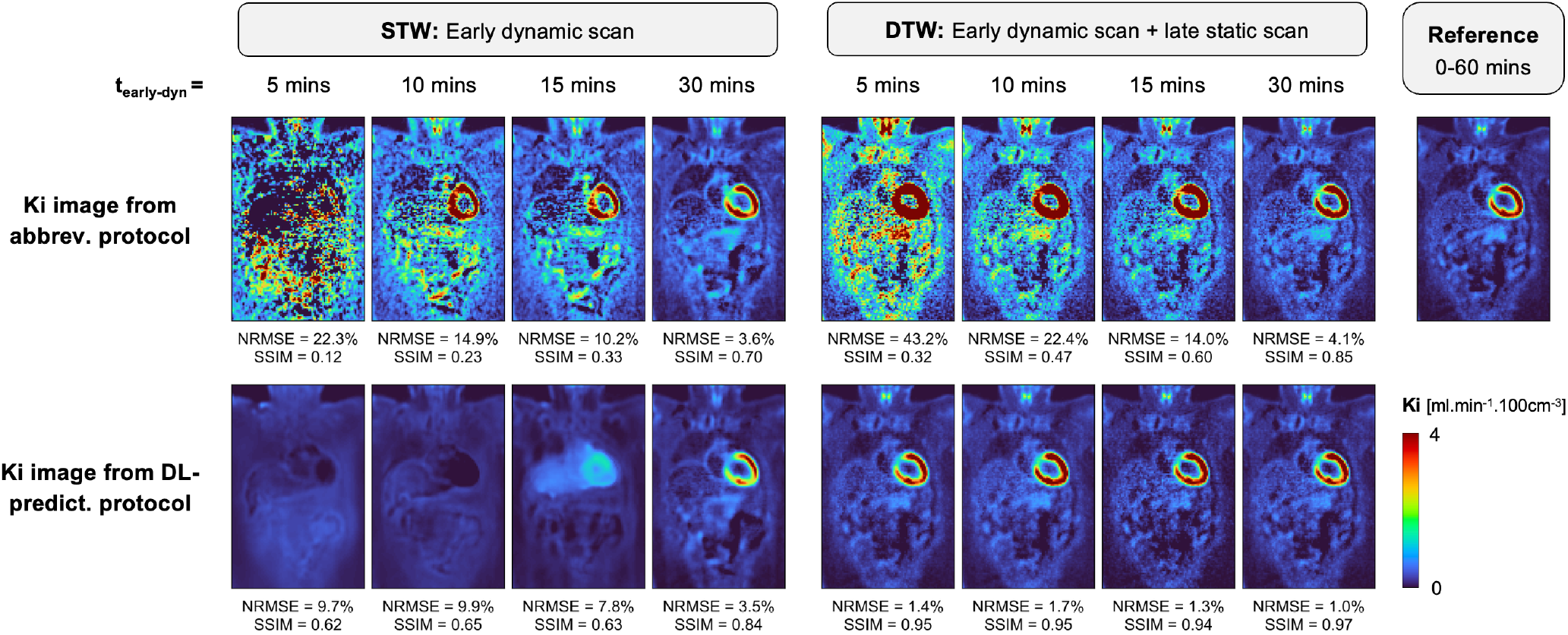
Coronal Patlak-derived K_i_ images of a test subject from the AUD/KE cohort. Columns compare STW and DTW protocols for t_early-dyn_ of 5, 10, 15, and 30 min. Top row shows FDG K_i_ images generated directly from the abbreviated (non-DL) dynamic protocol, while the bottom row shows K_i_ images derived from the DL-predicted frame sequences. The rightmost column shows the reference K_i_ image generated from the full-dose 0-60 min dynamic scan. For each protocol, the NRMSE(%) and SSIM values are reported with respect to the reference K_i_ image, with both metrics computed over the entire 3D imaging volume.

### B. Low-Dose Protocols

The above results for protocols at full-dose showed that less than 10 min of early dynamic data is insufficient for robust kinetic quantification using either STW or DTW with DL. Thus, the dose-level analyses in **Fig. 8** and **Fig. 10** focus on protocols at t_early-dyn_ = 15 min, while **Fig. 9** presents trends for t_early-dyn_ ≥ 10 min. **Fig. 8** compares the 0-60 min TACs with those from STW + DL and DTW + DL across five dose levels. Only the muscle and the two breast lesion phenotypes are shown, as these were the most challenging regions with the largest differences between STW and DTW at full dose (**Fig. 5**). As dose decreased, reference TACs became increasingly noisy, particularly in the muscle, where pronounced fluctuations emerged that were absent at full dose. In contrast, DL preserved smoother TACs that more closely reflected the expected physiological kinetics. With DTW + DL, ΔAUC remained within 3% for muscle and 10% for breast lesions across all dose levels. STW + DL showed larger deviations at lower doses, muscle ΔAUC increased from –9.7% (full-dose) to –15.7% (1/20 dose), while breast lesion ΔAUC exceeded 20% at all reduced dose levels. **Fig. 9** summarizes the mean differences in FDG K_i_ across all test subjects, calculated as the abbreviated protocol K_i_ minus the corresponding full-dose 60-min reference K_i_, for each dose level and t_early-dyn_ of 10-30 mins. K_i_ differences increased with lower doses, particularly below 1/5 dose. STW generally showed the largest deviations, especially in the liver, muscle, and breast lesion. Incorporating DL in STW reduced these differences except in the breast lesion. DTW produced smaller K_i_ differences overall, with DTW + DL providing the most stable estimates and the least dose dependence. **Fig. 10** compares representative K_i_ parametric images using Patlak fitting from the reference and abbreviated protocols at t_early-dyn_ = 15 mins. Dose reduction increased noise and resulted in pronounced overestimation of brain net influx values in STW and DTW protocols without DL. The noise was markedly suppressed in all DL-predicted sequences. However, STW + DL continued to exhibit unreliable flux distributions and artifacts, as also observed in **Fig. 7**. In comparison, DTW + DL produced the closest visual agreement with the reference, remaining robust across dose levels above 1/10 dose (37 MBq).

**Fig. 8:**
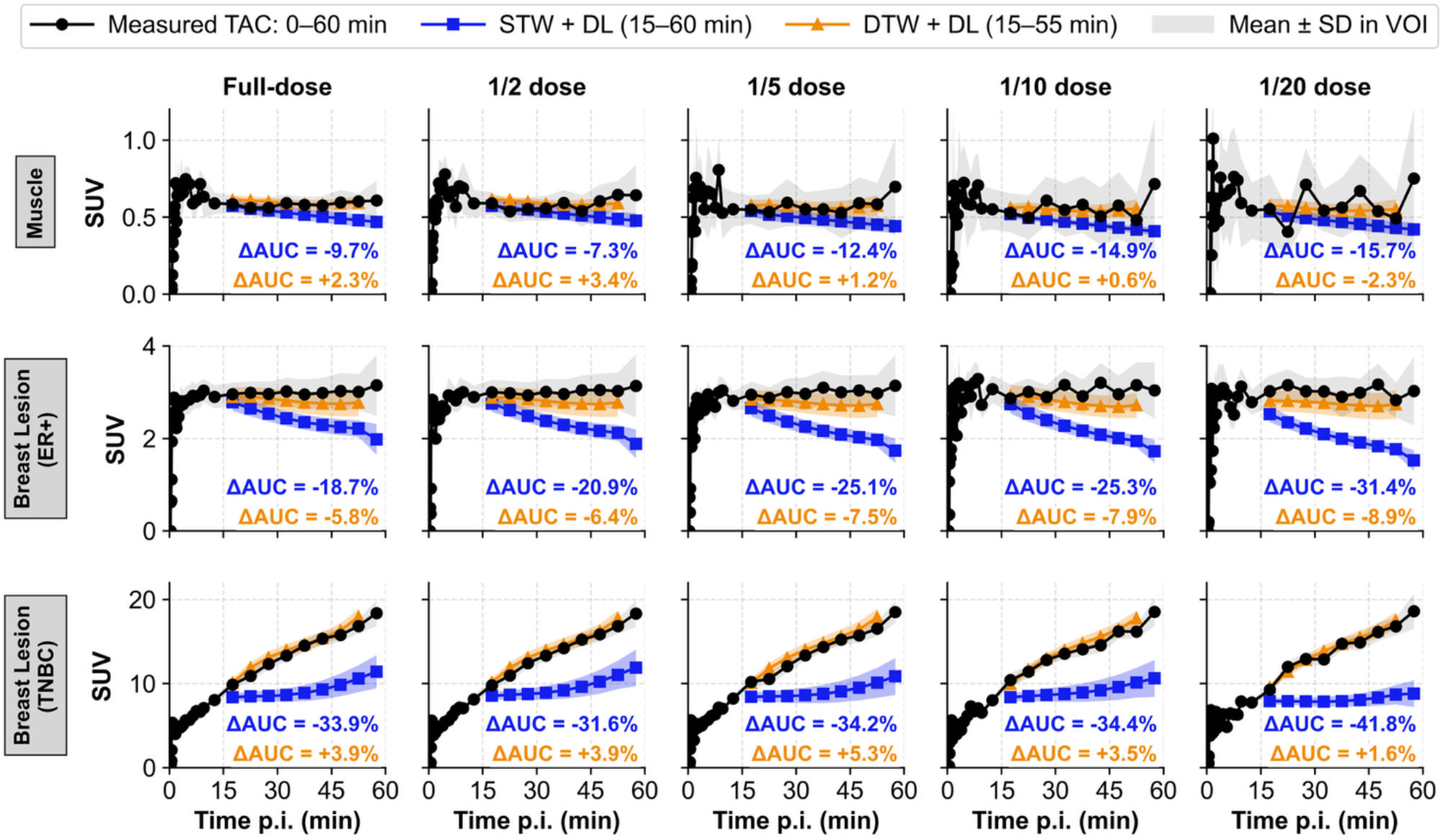
Representative TACs extracted from the muscle (of a subject in the AUD/KE cohort), an ER+ breast lesion, and a breast lesion from a TNBC subject compared across five dose levels. Reference TACs (0-60 min) are shown in black. TACs using STW + DL (predicting 15-60 min) and DTW + DL (predicting 15-55 min) are shown in blue and orange, respectively. Percent differences in area under the curve (ΔAUC) relative to the reference are reported.

**Fig. 9:**
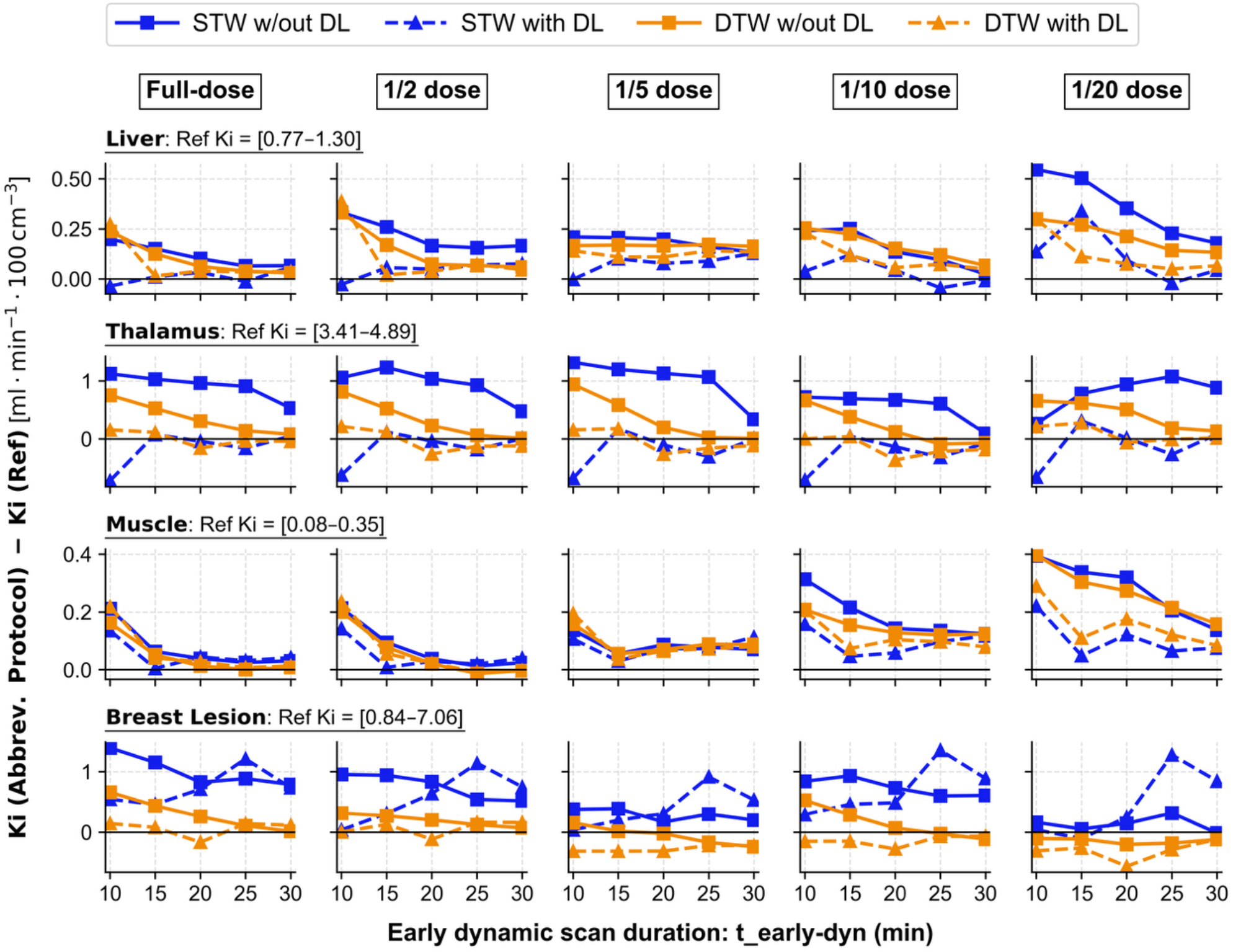
Mean difference in FDG K_i_ values estimated using 2-TCM fitting relative to the 60-min dynamic reference scan for the liver, thalamus, muscle, and breast lesion. Results are averaged across six test subjects, except for the breast lesion VOI, which includes four test subjects. Columns represent five dose levels. Results are shown for the STW and DTW protocols with and without DL. K_i_ values displayed in the subtitles indicate the range (min-max) of reference K_i_ values from the 60-min scan across testing subjects for a particular region of interest.

**Fig. 10:**
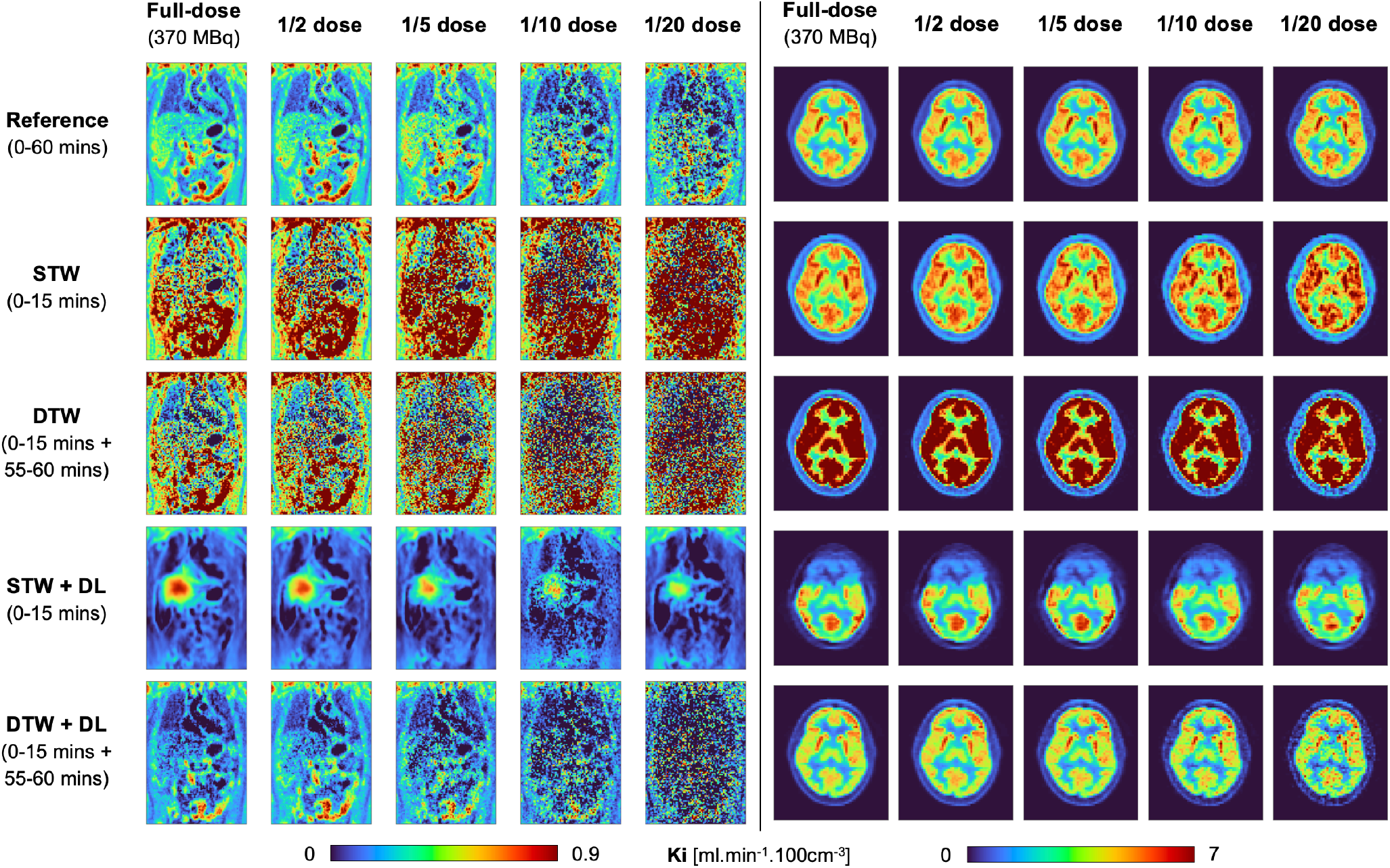
Representative Patlak K_i_ parametric images of the thoracic-abdominal region (left) and brain (right) compared between the reference protocol (0-60 min), STW (t_early-dyn_ = 15 min), DTW (t_early-dyn_ = 15 min + 55-60 min), and their corresponding DL-predicted sequences. Columns show results at five different dose levels.

## IV. Discussion

In this work, spatiotemporal neural networks were trained to predict missing dynamic frames from abbreviated LAFOV [^18^F]FDG PET scans, comparing single- and dual-time-window protocols for different early scan durations. This study extends prior work on DL-enabled abbreviated dynamic LAFOV PET in three ways. First, we incorporated a staggered FALCON MoCorr preprocessing pipeline. Previous studies either neglected patient motion and acknowledged it as a limitation [25-27, 29-32], or performed image registration without evaluating its impact [28]. Second, we assessed our networks across the full quantitative workflow, from DL-predicted frames to VOI-based kinetic modeling and voxel-wise parametric imaging. Third, we evaluated the combined effects of scan abbreviation and dose reduction to identify the shortest, lowest-dose protocol that maintained quantitative accuracy.

### A. Impact of Abbreviating Scan Duration

Across all qualitative and quantitative metrics, prediction quality improved with longer t_early-dyn_, reflecting the additional temporal information available to the DL model. This trend was consistent across all tissues and agrees with previous studies reporting reduced performance with shorter protocols [19, 21, 28]. Although DL predictions generally reduced K_i_ bias, substantial instability persisted for t_early-dyn_ = 5-10 min, with fluctuating VOI K_i_ estimates and Patlak K_i_ maps showing anatomically implausible flux distributions. This protocol-dependent degradation below 10 min, even with DL prediction, has not been previously reported. Prior work either evaluated a single abbreviated protocol [25, 26], omitted kinetic quantification [27], or investigated K_i_ image quality only for t_early-dyn_ ≥10 min [28]. These instabilities likely reflect the limited information available from the early uptake phase, before equilibrium is reached, to predict later tracer kinetics. Overall, the results indicate that <10 min of early-phase data is insufficient for robust prediction across the full kinetic analysis pipeline, for both STW and DTW.

### B. Impact of Protocol: STW vs. DTW

A consistent result across all evaluation metrics was the superior performance of DL-based predictions for DTW over STW protocols, in line with the findings of Ding [28]. The inclusion of the late 55-60 min p.i. image constrains the network during the late phase of the TAC. In contrast, STW protocols require the DL model to infer 30-55 min of tracer kinetics from early data alone, allowing autoregressive errors to accumulate. This limitation was most evident for breast lesions: STW models predicted declining late-phase activity, reflecting behavior learned from normal tissues (i.e., the models were not trained on subjects with breast lesions), whereas reference TACs showed stable or increasing uptake. Prediction accuracy depended strongly on tissue type. Tissues whose kinetics are characterized during the initial uptake, such as the lung, were predicted accurately with both abbreviated protocols. In contrast, lesion TACs differed according to phenotype. The ER+ lesion showed relatively low and stable late uptake, whereas the TNBC lesion exhibited continuously increasing FDG uptake. In these cases, including the late frame in the DTW protocol substantially improved TAC prediction by constraining the lesion’s late kinetic behavior.

Our results support a DTW protocol comprising a 0-15 min dynamic scan followed by a 55-60 min static scan with DL-based frame prediction. This agrees with Viswanath et al. [21], who reported that a 0-15 min plus 60-65 min protocol achieved FDG K_i_ bias within ±10% for high-flux breast tumors without deep learning. However, their analysis focused only on breast lesions, whereas the present study evaluated multiple organs and tissue types with diverse kinetic characteristics, providing a broader assessment of LAFOV PET protocol abbreviation.

### C. Impact of Reducing Dose

The DL-based frame prediction model remained largely robust to dose reduction. Training on mixed-dose abbreviated input sequences, each mapped to full-dose 60-min targets, gave the network an implicit denoising effect. For the DTW + DL protocol, deviations in tissue K_i_ values became evident only at 1/10 and 1/20 dose, where parametric images also exhibited increased noise, suggesting that reliable performance was maintained down to 1/5 dose (∼ 70 MBq). The degradation observed at very low doses is unlikely to be mitigated by extending the early scan duration alone, as the driving factor is reduced count statistics compromising the early frames used for prediction. Our previous work [36] showed that DL-based denoising improves kinetic parameter estimation for low-dose dynamic scans, including dual-tracer protocols. Ongoing work aims to first denoise the acquired frames and then use them to predict the temporal behavior of the missing data.

While the higher sensitivity of LAFOV PET enables abbreviated protocols to be combined with dose reduction (here: 5-fold), similar abbreviated acquisition strategies could potentially be translated to SAFOV systems, although without a comparable reduction in administered dose. Notably, LAFOV PET offers dose reduction while also enabling multi-organ imaging with measurement of the blood IF.

### D. Workflow Implications of Abbreviated Protocols

Both STW and DTW protocols shorten in-scanner acquisition sessions, decreasing the likelihood of within-scan motion and improving patient comfort and compliance, particularly for subjects with claustrophobia or difficulty remaining still. We found that a DTW protocol comprising a 0-15 min early scan and a late 5-min static scan, combined with DL-based frame prediction, was the shortest protocol that maintained robust [^18^F]FDG kinetic quantification. Although Ding et al. [28] recommended 0-10 min early scan with 60-65 min late scan, our results indicated that extending the early scan to 15 min improved quantification in tissues with slower kinetics (e.g., muscle) and in lesions of different phenotypes (e.g., breast lesions of ER+ and TNBC). It also resulted in more reliable voxel-wise parametric imaging. As summarized in **Table I**, protocol recommendations depend on the tissue and should be tailored for a clinical question rather than “one-fits-all”.

While DTW protocols improve kinetic parameter estimation compared with STW, several practical challenges should be considered. The main limitation is the need for two imaging sessions, requiring additional patient handling and potentially complicating scheduling. Accurate registration between early dynamic and late static PET acquisitions is essential. Whether the staggered FALCON MoCorr strategy can directly be applied to abbreviated protocols remains to be established, as registration between separate imaging sessions is more challenging due to patient repositioning (i.e., getting on and off the bed). Reference-frame selection may also require optimization, as it has been shown to influence MoCorr performance [46]. A further consideration is attenuation correction, as a second CT for the late static SUV scan would increase radiation exposure. However, this can be mitigated using emerging DL-based attenuation correction methods, such as pseudo-CT generation from PET data [47]. The first CT could be registered to the pseudo-CT generated from the second PET scan, followed by PET frame registration and MoCorr after image reconstruction.

### E. Limitations and Future Work

First, the implemented MoCorr preprocessing pipeline has several limitations. MoCorr accuracy could not be validated against a ground-truth motion signal (e.g., optical motion tracking, simulated deformation fields), so the evaluations presented in **Supplemental Material** relied on comparative measures. Furthermore, the staggered pipeline is semi-automatic and requires subject-specific inspection, limiting scalability. Additionally, because MoCorr was applied after PET image reconstruction, the attenuation correction may be inaccurate, particularly at the lung-liver boundary or for large gross body motions. Ideally, the estimated deformation fields would be used to update the attenuation image and then re-reconstruct the PET data. Lastly, MoCorr was performed on full-dose images, with deformation fields transferred to the low-dose images. Whether comparable performance can be achieved when MoCorr is performed on low-dose dynamic data remains to be investigated.

Our network shares conceptual similarities with the model by Ding et al. [28], which also exploits the spatiotemporal structure of dynamic PET data using convolutional LSTM, with additional recurrent units proposed in their framework. Their bidirectional prediction scheme incorporates both forward and backward temporal information but restricts prediction to fixed scan durations (65 min total) [28]. In contrast, our framework operates autoregressively in the forward direction and is thus not inherently limited to predicting time points seen in the training. Because each predicted frame is recursively fed back as input, the network can, in principle, generate arbitrarily long sequences, but prediction errors inevitably accumulate with successive frames. This behavior was not investigated here and warrants future study.

Another limitation is that all experiments exclusively used [^18^F]FDG data. Since dynamic imaging studies inherently depend on tracer kinetics, the present findings are specific for FDG and should not be assumed to predict performance for other tracers. Incorporating tracer-specific modeling into the network [26, 30, 32] could support extension to other tracers, including sequential dual-tracer imaging protocols. Finally, all quantitative analyses were restricted to the net influx rate K_i_. Extending the evaluation to other kinetic parameters (e.g., K_1_, k_2_, k_3_) represents an important direction for future work.

## V. Conclusions

In this work, we investigated the potential of DL to enable abbreviated dynamic [^18^F]FDG PET on the PennPET Explorer across multiple dose levels. A spatiotemporal ConvLSTM-based network was developed to autoregressively predict missing dynamic PET frames from abbreviated acquisition protocols. The recommended protocol is a dual-time-window (DTW) comprising a 15-min early dynamic scan followed by a 5-min late static scan, although tissue-specific differences suggest that protocol optimization is tissue-dependent and should be tailored to the clinical application. Combined with DL, this protocol achieved mean absolute FDG K_i_differences <10% in the liver and thalamus and approximately 10% in breast lesions, while the muscle remained the most challenging tissue (15.5%). Relative to the 60-min reference, the DTW + DL also reduced inter-subject variability compared with both the non-DL and STW + DL approaches while improving the quality of Patlak-derived K_i_ maps. Training on mixed-dose inputs and full-dose targets also imparted a denoising effect, enabling robust kinetic quantification up to a 5-fold reduction in dose (∼74 MBq), which is particularly relevant for future dual-tracer studies using same-day sequential injections. Future work should focus on multi-parametric and cross-tracer evaluation and extend the DL model development with dedicated denoising strategies and kinetics-informed learning.

## Data Availability

All data produced in the present study are available upon reasonable request to the authors.

## Acknowledgment

JC and FMM gratefully acknowledge Sara Neyt and Boris Vervenne (UGent) for help with PMOD and GPU machine, respectively. The authors thank Corinde Wiers (UPenn) for the generous use of her FDG data.

