## Supplemental Material for "Deep Learning Frame Prediction for Abbreviated Low-Dose Dynamic PET Protocols on the PennPET Explorer"

### S1. Additional Information: Preprocessing - Motion Correction

#### S1.1. Methods

Two FALCON-based preprocessing pipelines for MoCorr were implemented and compared (see **Fig. 2**): the *default FALCON* and a *staggered FALCON* method. The latter aims to address two limitations of the default FALCON pipeline, also acknowledged by Sundar *et al.* [23] in their original contribution: respiratory motion in the early dynamic frames and bladder-filling mismatch between early and late frames. Both approaches use the last frame (55-60 min p.i.) as the non-moving reference. The underlying registration algorithm within FALCON uses the normalized correlation coefficient (NCC), L-BFGS optimization, and Gaussian multi-scale framework.

- The **default FALCON** pipeline first registers frames 16-38 (1.5-55 mins) to the reference frame (frame 39, 55-60 mins), followed by a separate registration of frame 15 (80-90s) to the motion-corrected frame 16. The resulting deformation fields are propagated to all earlier dynamic frames (0-80s p.i.). While effective, this may introduce registration inaccuracies in the pelvic region due to mismatches in bladder-filling and residual motion in the lung-liver-kidney regions during early frames (50-90 s p.i.).
- The **staggered FALCON** pipeline groups dynamic image frames into sequential batches with comparable tracer distributions. Starting from last frame (55-60 min p.i.) as the fixed reference, registration is performed within each batch, with each newly motion-corrected frame serving as the reference for the preceding frame in the next batch. For the early dynamic frames (50-90s p.i.), each frame is treated as an individual batch

*Evaluation:* The performance of the MoCorr framework was evaluated by comparing three preprocessing strategies: no MoCorr, the default FALCON, and staggered FALCON. We note that no ground-truth motion information was available to assess the absolute accuracy of the estimated motion fields. Since both pipelines were based on the previously validated FALCON framework [23], our evaluation focused on a relative comparison between the default and staggered FALCON. Qualitative assessment evaluated the ability of the staggered FALCON to mitigate motion in the bladder region and the lung-liver-kidney region during the early dynamic frames. Gross body motion was visualized using *penumbra images*, generated by averaging voxel-wise binary body masks (SUV > 10% of the frame mean SUV) across all dynamic frames. In the resulting penumbra images, voxel values close to 1 indicated consistent inclusion within the body mask, values close to 0 indicated exclusion, and intermediate values (0.3-0.7) represented penumbra regions near body contours, with narrower regions indicating reduced inter-frame motion. Quantitative performance was evaluated in the 13 subjects from the AUD/KE study using the normalized correlation coefficient (NCC). For each pair of consecutive frames, voxel-wise NCC maps were computed using 4x4x4 voxel patches, and the mean NCC was calculated for each map.

#### S1.2. Results

**Fig. S1** and **S2** present qualitative comparisons to show the improvements provided by the staggered FALCON approach to address registration artifacts from bladder filling mismatch and involuntary lung-liver-kidney motion in the early frames. In **Fig. S1**, maximum intensity projection (MIP) images are shown for the 180-210s p.i. frame, comparing motion-corrected images with default and staggered FALCON. The red arrows in the default FALCON image indicate registration artifacts in the vasculature. These artifacts arise from mismatches in bladder filling between moving and reference frame, which are reduced with the staggered FALCON. In **Fig. 2**, coronal image frames without MoCorr illustrate involuntary motion across early dynamic frames (here shown for 45-80s p.i.) in the lung-liver-kidney region. Using the overlaid horizontal red dashed lines as a visual reference, it can be observed that the kidney boundaries and liver-lung interface are inconsistently aligned across frames. To assess the MoCorr applied by the default and staggered FALCON, difference images relative to the corresponding non-motion-corrected images are shown in the bottom two rows of **Fig. S2**. Because these images represent the voxel-wise differences relative to the non-motion-corrected images, larger differences reflect larger applied deformations and therefore greater MoCorr. Compared with the default FALCON, the staggered FALCON produces larger differences along organ boundaries, particularly in the right kidney, indicating more extensive correction of motion. For regions where motion was originally already limited (e.g., spine, lung), the staggered FALCON shows less pronounced differences than the default, suggesting that unnecessary deformations are avoided.

**Fig. S3** shows penumbra images to visualize the correction of gross body motion. With no MoCorr (left column), pronounced penumbra regions are present along the arms, the back, and legs (indicated in the white boxes). In contrast, penumbra images for both the default and staggered FALCON show a substantial reduction in these regions, indicating comparable correction of motion at the body contours. This similarity is expected, since the staggered FALCON primarily addresses involuntary liver-lung-kidney motion and bladder-filling mismatches rather than gross body motion.

**Fig. S4** compares the frame-by-frame NCC for no MoCorr, the default FALCON, and staggered FALCON. Both methods consistently yield higher NCC values than no MoCorr, with staggered FALCON achieving the highest values (33% improvement in NCC compared to a 24% increase for default FALCON). This improvement is primarily due to the staggered FALCON correcting lung-liver-kidney motion from frames 10-15 (50-90s p.i.) onward and resolving the bladder-filling mismatch observed with the default FALCON. Note that because no explicit MoCorr is applied during the first 10 frames (< 50s p.i.), the modest increase in NCC observed during this period is unlikely to reflect improved motion compensation; instead, it likely arises from smoothing introduced by interpolations during deformation field resampling.

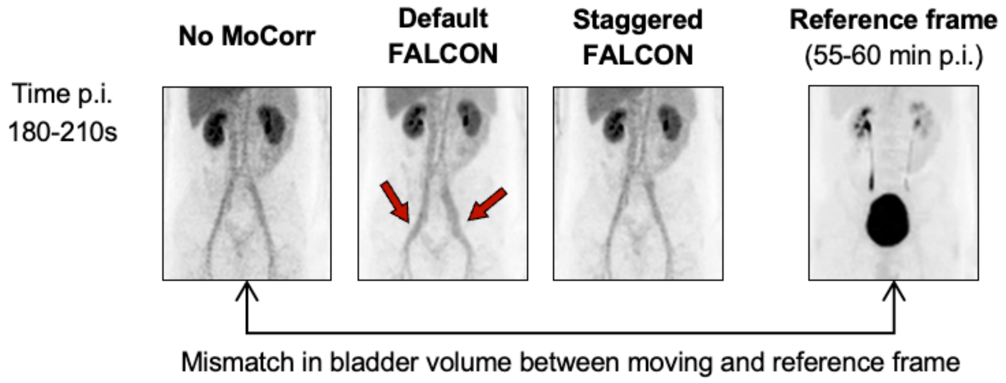


**Fig. S1:** Qualitative MoCorr results. Coronal MIP images (180-210s p.i.) showing the artifact in the bladder vasculature (arrows) with default FALCON and its correction with staggered FALCON. The reference frame (55-60 mins p.i.) is included to illustrate the bladder filling mismatch.

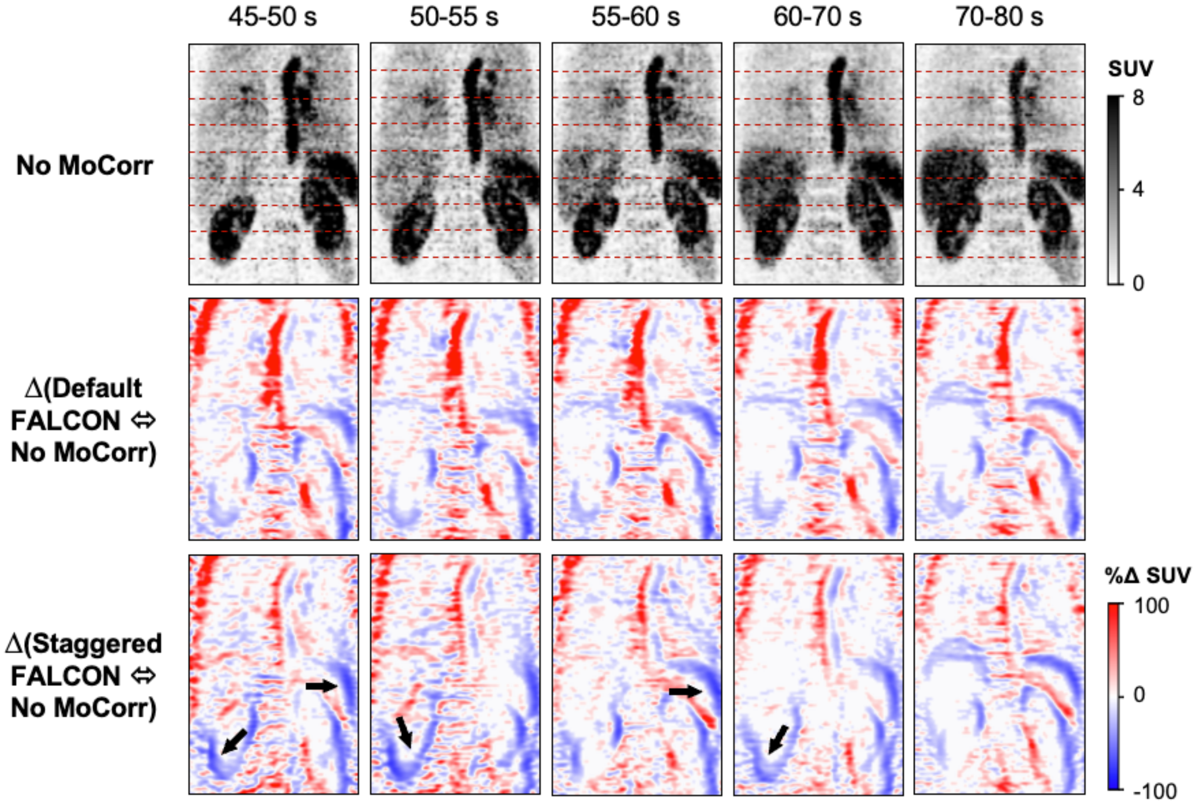


**Fig. S2:** Qualitative MoCorr results. Coronal images of the early dynamic frames (45-80 s p.i.). The top row shows no MoCorr; the bottom rows show voxel-wise difference images relative to no MoCorr for default and staggered FALCON, where larger differences indicate larger applied deformations.

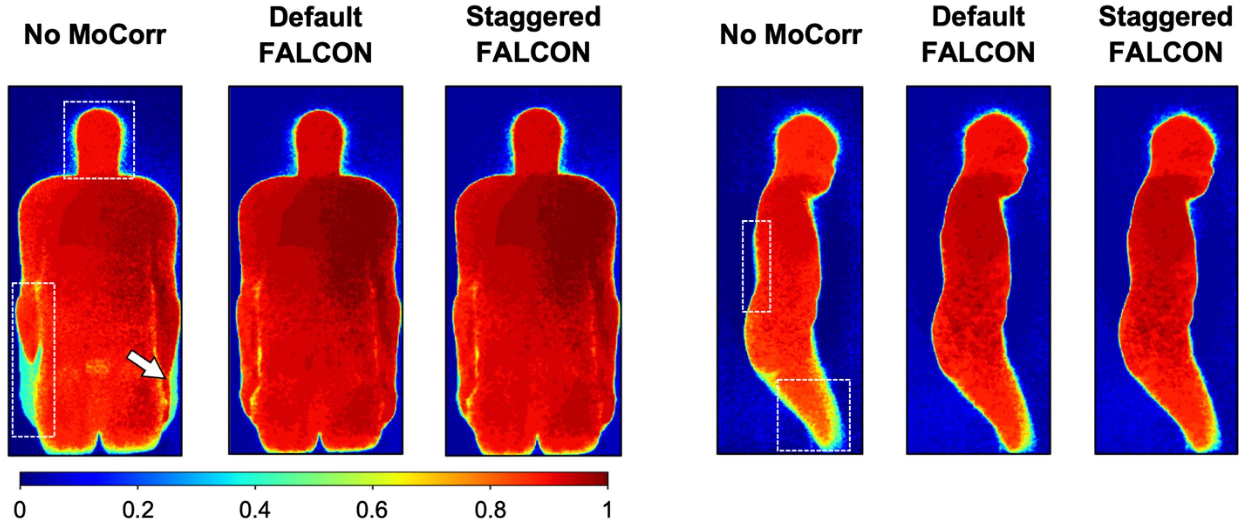


**Fig. S3:** Penumbra images, computed as the voxel-wise mean of the body masks across all dynamic frames. Values range from 0 (always outside the body mask) to 1 (always inside), while intermediate values represent penumbra regions corresponding to body contours affected by gross patient motion. Dashed boxes and arrow highlight representative regions.

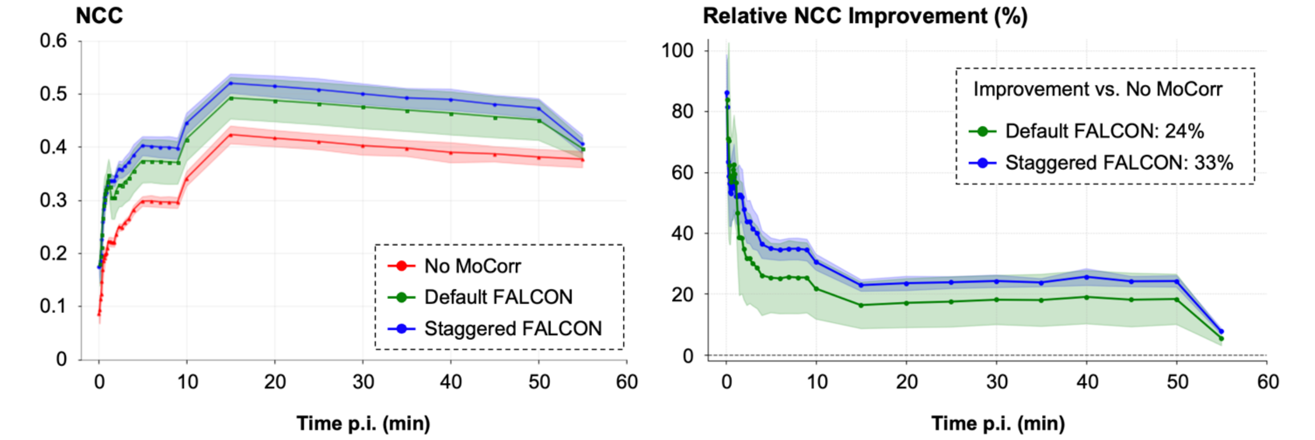


**Fig. S4:** Quantitative MoCorr results, comparing no MoCorr, the default FALCON and staggered FALCON. **Left:** Frame-by-frame normalized correlation coefficient (NCC), averaged across the 13 subjects from the AUD/KE study, shown as a function of frame timings. **Right:** Relative NCC improvement (%) of default FALCON and staggered FALCON compared with no MoCorr. The values in the text box (default: 24%; staggered: 33%) represent the average improvements calculated over frames acquired after 1.5 min p.i.
